# MRI-Based Multi-Class Relevance Vector Machine Classification of Neurodegenerative Diseases

**DOI:** 10.1101/2024.10.07.24315054

**Authors:** Kyan Younes, Yann Cobigo, Amy Wolf, John Kornak, Katherine P. Rankin, Mirza Faisal Beg, Lei Wang, Howard J. Rosen

## Abstract

Machine learning algorithms are a promising automated candidate that can help mitigate the growing need for dementia experts. Despite the substantial development in MRI-based machine learning analyses, case misclassification is a universal finding, yet the reasons behind misclassification are poorly understood. We implemented a multi-class classification approach that uses relevance vector machine and logistic classification to classify research participants based on their whole-brain T1-weighted MRI scans. A total of 468 participants from seven diagnostic classes were included: 144 healthy controls, 84 Alzheimer’s disease, 108 behavioral variant frontotemporal dementia (bvFTD), 30 semantic variant primary progressive aphasia (svPPA), 30 non-fluent variant primary progressive aphasia (nfvPPA), 30 corticobasal syndrome (CBS), and 42 progressive supranuclear palsy syndrome (PSPS). We compared the algorithm‘s diagnostic accuracy against the clinical, pathological, genetic, and quantitative imaging data. The exact neurodegenerative syndrome was predicted in 71% of the cases, the neurodegenerative disease spectrum was predicted in 80% of the cases, and the algorithm distinguished controls from any dementia in 85% of the cases. The algorithm showed high performance in diagnosing healthy controls, moderate performance in diagnosing AD, bvFTD, and svPPA, and low performance in diagnosing CBS, nfvPPA, and PSPS. Based on the quantitative imaging data, most of the misclassified neurodegenerative cases had minimal atrophy and brain volumes comparable to healthy controls. In AD, early-onset AD cases with minimal brain atrophy represented most of the misclassified cases. In bvFTD, FTD genetic mutation carriers (predominantly *C9orf72* repeat expansion), FTD phenocopy, patients meeting only possible bvFTD criteria represented most misclassified cases. Case misclassification in machine learning studies in neurodegenerative diseases results from neurodegenerative disease heterogeneity and the limitations of structural MRI’s ability to capture the whole gamut of biological changes. Larger and more inclusive datasets that are representative of population biologic heterogeneity are needed to train better machine learning techniques, and a margin of error is expected and should be acceptable, like the uncertainty of a clinical diagnosis by a dementia expert.

## Introduction

Around 50 million people have dementia worldwide, and there are nearly 10 million new cases diagnosed annually [1]. These growing numbers represent a global challenge with catastrophic humanitarian and economic consequences [2]. While Alzheimer’s disease (AD) is the most common cause of dementia, there is a number of other neurodegenerative diseases that lead to dementia, and these become increasingly more prevelant in patients with young-onset neurodegnerative disease. Frontotemporal dementia (FTD), in particular, is about as common as Alzheimer’s disease in those whose onset of dementia occurs before age 65 [3]. Accurate diagnosis of patients with behavoiral or cognitive symptoms is critical for proper triaging, developing care plan, treatment, and referral to theraputic clinical trials. Dementia diagnosis is, however, a labor-intensive process that often requires a multi-disciplinary team of specialists, almost exclusively available at referral centers [4, 5]. Brain imaging is an important part of the diagnostic process, helping to confirm that neurodegeneration is taking place and helping to predict a specific diagnosis [6, 7, 8]. Despite the prominence of neuroimaging in the assessment of patients with cognitive complaints and the large array of methodologies used to quantify abnormalities, visual interpretation remains the mainstay in evaluating dementia patients in clinics. This means that the diagnosis depends on unique skills and experience with image interpretation that can vary even across experts. Automated machine learning algorithms have the potential for making accurate diagnostic classifications based on brain imaging, which can potentially improve the reproducibility and accuracy of dementia diagnosis and create opportunities for accurate diagnosis even in settings where expert physicians are not available. However, many limitations, including unexplained misclassifications, continue to hinder the widespread use of machine learning algorithms for the diagnosis of neurodegenerative diseases [9, 10].

Many machine learning algorithms have been studied in neurodegenerative illnesses [9, 10]. One practical limitation of many of these studies is the use of binary classification (e.g. distinction of AD from FTD) [11, 12, 13, öller2016**?**, 14, 15, 16]. In clinical practice, although a single diagnosis is sought after, clinical presentations do not always conform to the features highlighted in published diagnostic criteria, and symptoms can change over time as disease progresses [4][17]. Clinical judgment must account for these complexities, and dementia specialists naturally generate differential diagnoses that addresses multiple possible causes rather than choosing between binary possibilities. Recognizing the limitations of binary classification, few studies have implemented machine learning algorithms that attempt to differentiate among a variety of major neurodegenerative illnesses including AD, FTD, Lewy body dementia (LBD), vascular dementia, and primary progressive aphasia (PPA) [18, 19, 20, 21, 10, 9]. Most of the studies that examined FTD included behavioral variant FTD (bvFTD) and PPA variants as one diagnostic group (*i.e.,* FTD), which can inflate the accuracy of the results because certain variants, such as semantic variant (svPPA), tend to have unique distinguishing features that permit higher diagnostic accuracy compared with other variants of FTD [22, 19, 20, 23, 24, 25, 18]. When classifying among the FTD variants, studies either included only PPA variants without bvFTD [26, 27] or did not differentiate bvFTD and PPA variants from healthy controls and AD [21]. Additionally, there have been no studies that included two important sub-types of the FTD-spectrum disorders; namely: cortiobasal syndrome (CBS) and progressive supranuclear palsy syndrome (PSPS), where symptoms can overlap with those of bvFTD and PPA.

Some of the best-performing multi-class machine learning techniques in neurodegeneretative disease classification implement support vector machines (SVM) [9, 10, 28]. However no studies, to our knowledge, have used relevance vector machines (RVM) [29]. RVM is similar to SVM, except that it implements a Bayesian framework, provides a probability distribution, and produces a smaller set of vectors to discriminate between groups [30, 31, 32]. Whereas SVM separates groups by placing support vectors close to the boundaries of the hyperplane that discriminates the groups, RVM places relevance vectors deep within each group leading to higher specificity [33, 29]. We speculate that RVM has advantages over SVM given the relatively low number of neuroimaging features typically required by a dementia specialist to generate a differential diagnosis [34, 35, 36]. Another limitation to the current multi-class classifiers is that they base the results on the class with the highest probability rather than using the relative distribution for each of the classes within a probability distribution. In an effort to use all the data provided by the probability distribution, we implemented a posterior logistic classification on the RVM output. Lastly, case misclassification is common across machine learning studies in neurodegenerative illnesses [10, 9], yet few studies seek to explore the reasons behind case misclassification.

In this study, we examined the performance of an RVM-based classification for neurodegenerative diseases using a data-driven, multi-class classification algorithm and posterior logistic classification tuning on T1-weighted whole-brain MRI with expert clinical diagnosis as the gold standard. To emulate the probabilistic approach a dementia specialist employs to generate a differential diagnosis, we employed a probabilistic classification across seven neurodegenerative disease classes: AD, the full clinical range of FTD spectrum disorders (*i.e.,* bvFTD, svPPA, nonfluent agramatical PPA (nfvPPA), CBS, and PSPS), and healthy controls. We further examined our results in a subset of the study participants who had available pathology and genetic data. Lastly, we also examined quantitative neuroimaging, genetic, and pathological data to provide insights into the features that lead to accurate versus inaccurate predictions.

## Methods

### Subjects Selection

Participants of this study (N=468) were recruited and assessed at the University of California, San Francisco (UCSF), Memory and Aging Center (MAC) through several projects funded to study AD and FTD. Diagnosis for these studies was made per published criteria based on a multidisciplinary evaluation and a consensus conference that included behavioral neurology, neuropsychology, and nursing input [5]. Typically, patients were followed for at least two consecutive years. To maximize the value of the classification system for clinical diagnosis, we used MRI data from the first time point for each participant. However, to ensure optimal algorithm training, a gold standard clinical diagnosis for each case was established after considering all of the clinical data available from all the research visits for each participant. We included seven diagnostic classes; AD, bvFTD, nfvPPA, svPPA, CBS, PSPS, and cognitively healthy control (CO). Because AD and FTD-spectrum prevalence are roughly equal in prevalence in those with early age of onset dementia [37], we limited our analysis to patients younger than 70 years old, where the likelihood of FTD-spectrum disorders is highest, so that the RVM would be trained on a clinical dataset relevant to the clinical decisions made in practice. We included cases that showed diagnostic stability overtime even when the diagnostic certainty was low such as possible bvFTD and FTD-phenocopy [38, **?**]. However, we excluded patients whose clinical syndromic diagnosis changed over the years, as this has been associated with lower diagnostic accuracy. We excluded patients with vascular disease-only, and those with mild cognitive impairment (MCI) that did not go on to develop dementia syndrome. However, they were included if the diagnoses changed subsequently to one of the syndromes included in this study. Patients who had evidence of vascular disease in addition to any of the neurodegenerative syndromes were included. Patients with FTD/motor neuron disease (MND) were included, but patients with MND-only were excluded. We did not include patients with Lewy body dementia because too few cases were available.

CO data were obtained from the Hillblom Healthy Aging Study [39]. Participants are recruited via advertisements and community events. CO underwent the same evaluation as disease group patients and were required to have no clinically significant cognitive or behavioral complaints, performance within one standard deviation of normal on all cognitive tasks, and to have brought a knowledgeable informant to the visit to verify the absence of clinically significant cognitive or behavioral changes. Controls were excluded if they had a history of significant mood problems, clinically significant alcohol or drug use, significant vascular disease, visual problems that would impair test performance, other neurological conditions, and self-reported deficits in cognition.All research was performed in accordance with the Code of Ethics of the World Medical Association.

#### Genetic and Neuropathology Data

A subset of 417 patients (89% of the sample) had genetic data available for the following genes: *PGRN, MAPT, TARDBP, C9orf72, APP, PSEN1, PSEN2, FUS, APOE, and MAPT H1/H2* haplotypes. Seventy-two patients (15% of the total cohort, 22 percent of the disease cohort) had available pathology data. At autopsy, the brains were processed and analyzed according to the UCSF Neurodegenerative Disease Brain Bank protocol [40]. In short, 8*µ*m thick formalin-fixed paraffin-embedded sections from 27 regions of interest underwent routine staining and immunohistochemistry for hyperphosphorylated tau, amyloid-*β*, TDP-43, alpha-synuclein, and 3R-tau antibodies. Neuropathological diagnoses were made based on consensus criteria [41, 42, 43].

### Image Acquisition

All subjects provided informed consent, and the clinical and imaging protocols were approved by the UCSF Committee on Human Research. Participants underwent a whole-brain magnetic resonance imaging (MRI) on a 3 Tesla Siemens Trio Total imaging matrix (Tim) scanner with a 12-channel head coil, or a Prisma Fit scanner with a 64-channel head coil (Siemens Medical Solutions ^1^, Erlangen, Germany). T1-weighted images were acquired with a Magnetization Prepared Rapid Gradient Echo (MP-RAGE) sequence (Sagittal slice orientation; 240 *×* 256 matrix; 160 slices; voxel size = 1.0 *×* 1.0 × 1.0 mm^3^; TR = 2300 ms; TE = 2.98 ms; flip angle = 9*^◦^*).

#### Image processing

Before any preprocessing, all T1-weighted images were visually inspected for quality. Images with excessive motion or other image artifacts were excluded. T1-weighted images underwent bias field correction using N3 algorithm [44, 45], and segmentation into gray matter (GM), white matter (WM), and Cerebrospinal fluid (CSF) tissue was performed using the SPM12 (Wellcome Trust Center for Neuroimaging ^2^, London, UK) unified segmentation [46]. A group template was generated from the segmented tissue probability maps by non-linear registration to a common space using the *Large Deformation Diffeomorphic Metric Mapping* framework [47]. Participant gray matter maps were normalized, modulated and smoothed in the group template space using a Gaussian kernel with 5 mm full width half maximum. Every transformation step of the transformation was carefully inspected from the native space to the group template.

### Classification Engine

The GM and WM voxels are first flattened into a column vector. However, the high voxel count can degrade the classification accuracy due to the inclusion of voxels that may contribute to over-fitting and, therefore, introduce noise. Therefore, we investigated various pre-classification methods to reduce the dimensionality of the input vectors. Pre-classification image treatments were conducted with Insight Toolkit (ITK) [48, 49], and linear algebra components were developed using Eigen3 [50]. We examined three different dimension reduction methods: 1) Voxel down-sampling, inside a tissue mask, using trilinear interpolation, with down-sampling factors of *f* = 2, 4, 8, 16. In other words, each voxel’s dimension is multiplied by the factor *f*. Here, we denote a down-sampling model with factor *f* as *fx*. For instance, the voxel of 1.5 × 1.5 × 1.5 mm^3^ in the *f* 2 model becomes 3 × 3 × 3 mm^3^ voxel. Three masks, thresholded at a tissue probability of 70% were used: the entire GM and WM (GM WM); the GM and midbrain, pons, superior cerebellar peduncle and thalamus (GM MPST) [51]; the GM and midbrain, superior cerebellar peduncle and thalamus (GM - MST). 2) Region of interest constrained GM and WM using the Neuromorphometrics atlas^3^ [52]. And 3) Principal Component Analysis (PCA) using singular value decomposition [53]. We considered a range of principal components: 0,2,3,4,5,10,15,20 and 50, with 0 representing the case without PCA dimension reduction. PCA was used as an additional dimension reduction layer after the implementation of method 1) or 2) [54]. In total, we tested 9 sets of PCAs for 4 down-sampling factors using 3 types of masks and one atlas: 9 × (4 × 3 + 1) models. In the results, *fx − y* represents the dimension reduction model with voxels down-sampled *x* times using *y* first PCAs, or *Atlas − y* refers to atlas-based downsampling using *y* first PCAs. As an example, the model *f* 2 *−* 20 represents an image downsampled by multiplying all the voxel dimensions by a factor *f* = 2 and using the 20 highest principal directions to build the feature vectors.

#### Relevance Vector Machine

The classification engine is based on the Relevance Vector Machine (RVM) [29] and was implemented in our framework using Dlib-ml [55]. RVM is a supervised learning algorithm often compared to Support Vector Machine (SVM) [56]. Like SVM, RVM is a binary (i.e., two-class) classification. The *n* input images are vectors 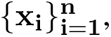 where **x_i_** = (**x_i1_**, **x_i2_***, · · ·,* **x_ip_**)**^T^** represents the vectorized GM and WM image for the subject *i* whose *p* predictors are the features described in the previous section. The symbol *T* represents the transposition of the vector. The labels 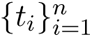 are the diagnoses attributed to each subject *i*. The decision function is 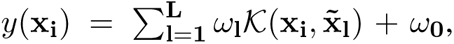 where *ω_l_* are the model fitted weights and 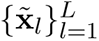 are the *L* relevance vectors [53]. The kernel function 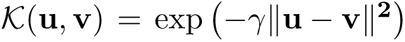 is the Radial Basis Function (RBF), where *γ* is the hyperparameter to fit. The RBF projects the data into a feature space emphasizing the similarity between the vectors **u** and **v**. The major difference between SVM and RVM resides in the under-jacent algorithm and the corresponding interpretation of the landmark vectors 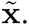 In SVM the landmarks are called support vectors and lie close to the margin delimiting the class boundaries. In RVM the landmark vectors are relevance vectors and represent the prototype vectors of each class and lie more deeply inside each class. Another fundamental difference is that the outputs are scores for SVM, whereas they are probabilities for RVM. However, it is possible to convert an SVM score into probabilities using Platt’s algorithm [57].

#### Multi-class classification

In this section, our objective is to construct a multi-class probability distribution over the training conditions. Each subject’s probability distribution represents the potential outcome according to a decision maker. Several algorithms extend two-class classification to *K*-class classification, where *K* represents the number of classes. A commonly used method is the voting scheme, which counts how many times a class wins in all *K*(*K −* 1)*/*2 pairwise comparisons, with the winning class being the one with the highest number of victories [58]. However, the voting scheme treats each decision function independently and does not account for the relative importance or competitiveness among classes. Consequently, in a scenario with *K* classes, the scheme processes all combinations of pairwise comparisons and simply selects the majority winner, potentially overlooking the nuanced relevance of other classes in the classification task. In our framework, we implemented the pairwise coupling algorithm proposed by Hastie and Tibshirani [59], which combines binary decision functions into a multi-class classification, resulting in a *K*-class probability distribution. In this approach, we let *r_ij_* = {*C* = *c_i_|X* = *x, c_i_ ∪ c_j_*}, *n_ij_*being the number of observations in the training binary set {*ij*}. Let the probabilities at feature vector **x** be **p**{**x**} = (*p*_1_(**x**)*, p*_2_(**x**)*, · · · p_K_*(**x**)). We wish to find *p_i_* so that the 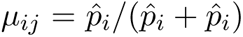 are close to the *r_ij_*’s. We have *K −* 1 free parameters: degrees of freedom. The minus one comes from the constraint 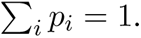 In order to find the best approximation *µ* Hastie and Tibshirani’s algorithm minimizes the average weighted Kullback-Leibler distance:

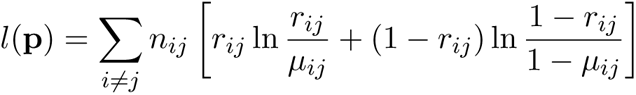

To determine the accuracy of the multi-class probability distributions as a diagnostic tool, the probability distributions were examined and compared to the clinical assessments.

### Nested Cross-Validation

Our study involved testing 9 × (4 × 3 + 1) dimension reduction models. To evaluate the best model, we implemented a nested cross-validation scenario with a *k_i_*-folds inner-loop and a *k_o_*-folds outer-loop, where *k_i_* and *k_o_* represent the number of folds in each level of the loops. Within the two nested loops, all diagnostic categories were proportionally divided into (*k −* 1)*/k* fractions for the training fold and 1*/k* fraction for the testing fold. Where *k* denotes the *k*_{_*_o,i_*_}_ of the described loops. For each training fold, two grid searches were performed to optimize the fitted hyper-parameters. A first search of 10^4^ parameters between 10*^−^*^7^ and 1, gave an approximate estimate of the hyper-parameter. A second, more refined grid search around the first estimate value examined 100 parameter choices within a range of 20% around the estimated value.

#### Selection of optimal parameters

To determine the best model we evaluated a total of 7 × (7 *−* 1)*/*2 combinations of binary classifications using previously described 9 ×(4 ×3 +1) configurations. This results in 2,457 models nested in a 6-folds outer-loop and having 3-folds inner-loop for each cross validation. For both inner- and the outer-loops we calculated the average accuracy (*Acc*) over the folds. A robust estimation of the performance was calculated by combining the mean and standard deviation of a metric *ν* into one estimator: *µ* = (*β − ν*) *× sd/ν*, where *sd* represents the standard deviation of *ν* over the folds, and *β* is the ultimate value *ν* can reach. The estimator *µ* can be interpreted as the inverse of the signal-to-noise ratio formula for *ν*, reinforced by the weight of the distance between *ν* and *β*: (*β − ν*). This approach allows us to avoid issues with null variance and places more emphasis on *ν*. We aimed for the smallest value of *µ*. In the following paragraphs, *ν* will be the accuracy of the binary classifications, with an ultimate value *β* = 1; and true positive rate (TPR) for the multiclass classification, with the ultimate value *β* = 1. The average of the TPR over all classes represents the multiclass balanced accuracy (b-acc). The *verification* set is defined as the set used to optimize the binary engines and reused to verify the pairwise coupling. The *validation* set is defined as the set left out from the outer-loop and used to validate the pairwise coupling. It is worth noting that the choice of the multiclass model is not based on the best performing binary classification. Instead, we selected the binary classification model that yielded the best results of the multiclass classification.

#### Logistic regression

The probability distribution resulting from our processing is a seven-dimensional feature space, which is difficult to visually interpret. Like the voting scheme, selecting the highest probability would result in an inadequate outcome, as it would ignore the information brought by the other classes and their combinations. We implemented a multiclass logistic classification, based on LogisticRegressionCV (form sklearn v0.20.4 python package) using 3-folds cross validation applied on the probability distributions of the verification dataset and calculated three accuracies: 1) the diagnostic accuracy (*ACC_dx_*) representing the accuracy based on each diagnosis; 2) the canonical accuracy (*ACC_canonical_*) representing the classification between controls, *AD* and *FTD*; and 3) the dementia accuracy (*ACC_dementia_*) representing the classification between the controls and all demented patients combined.

### Validating classification outcomes with the clinical and pathological data

For each case, the RVM outcome was compared with the clinical diagnosis, as well as the pathological diagnosis in the subgroup with available pathological data. Classification accuracy was evaluated on four levels: 1) *syndrome* – the exact clinical syndrome was predicted by RVM, *i.e.* AD vs. bvFTD vs. CBS vs. CO vs. nfvPPA vs. PSPS vs. svPPA; 2) *disease spectrum* – RVM predicted another clinical syndrome within the same disease spectrum, *i.e.,* AD vs FTD-spectrum vs. CO; 3) *disease* – RVM predicted a neurodegenerative disease but neither the exact syndrome nor within the same disease spectrum, *i.e.,* neurodegenerative vs. CO; 4) *wrong classification* – a neurodegenerative disease is classified as CO or a CO case is classified as a neurodegenerative disease. Each of these levels of classification represents a useful approach for clinical practice. For instance, even if the diagnostic process does not arrive at the exact diagnosis, raising a concern for an atypical disease (e.g., FTLD-spectrum) or a neurodegenerative disease in general is helpful clinically and can trigger further referrals and workup.

### Validating classification outcomes with W-score maps of the structural T1w images

To examine the imaging features associated with accurate or inaccurate classification, we quantified the degree of atrophy in each atlas ROI in each participant. W-score maps (W-score) were generated by comparing each patient’s gray matter maps to 534 neurologically healthy controls from the MAC Hillblom Healthy Aging Network (age range 44-99 years, M*±*SD: 68.7 *±*9.1; 220 male/302 female), adjusted for age, sex, total intracranial volume. Mean W-score values were extracted for each region of interest (ROI) in the probabilistic Desikan atlas.[60, 61] Given that regions of severe atrophy are generally presumed to be important contributors to classification, we examined the relationship between classification outcome and W-scores for the ROI’s with peak volume loss in each participant across the seven RVM output classes.

## Results

A total of 468 participants were included in the analysis. The demographic charachterestics of the cohort is shown in Table 1.

**Table 1:**
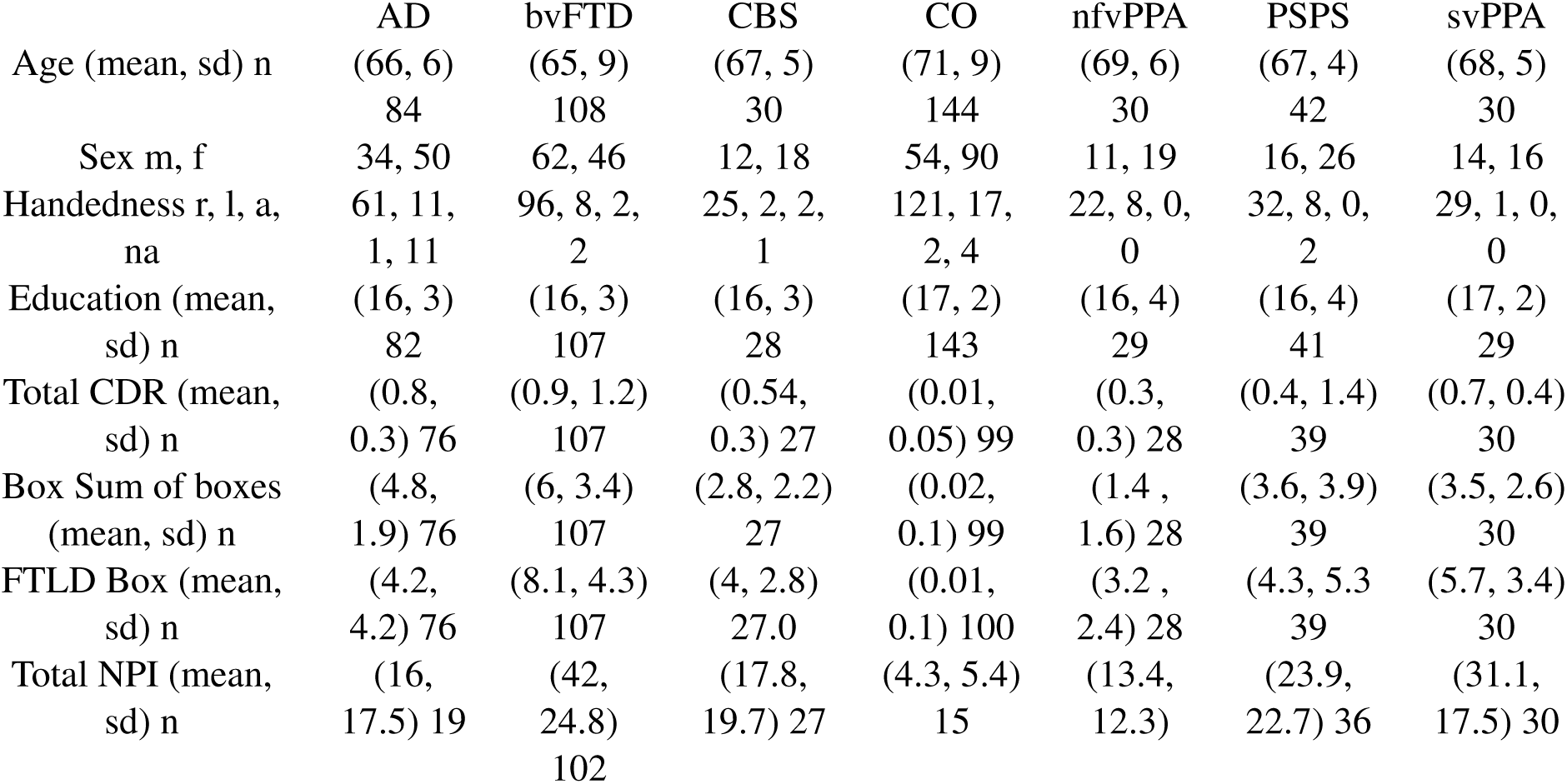
Cohort Demographics.

### Seven classes results

The focus of our study is on developing an efficient multi-class classification model. To that end, a summary of the highest-performing models is presented in Table 2, where the true positive rate (TPR) is derived from the confusion matrix and averaged across the seven validation folds. For illustration, an example confusion matrix is provided in Table 3. Figure 1 shows bvFTD’s probability distribution using the model *f* 2 *−* 20. The box plots represent the quartiles distributions for each outer-loop cross validation. When comparing the models in Table 2, it becomes evident that the best-performing models are voxel-wise, particularly those that employ smaller down-sampling and utilize the GM WM mask. A multiclass logistic regression was trained on the results of RVM and applied to the validation dataset, yielding an improved classifier specificity (Table 4) compared to the classifier without logistic regression correction (Table 3). Without the logistic correction, the accuracies were 0.69(0.06), 0.82(0.03), 0.85(0.03) for the diagnosis, canonical, and dementia groups, respectively. After applying the logistic regression correction, the accuracies improved to 0.72(0.06), 0.83(0.04), 0.86(0.04).

**Figure 1:**
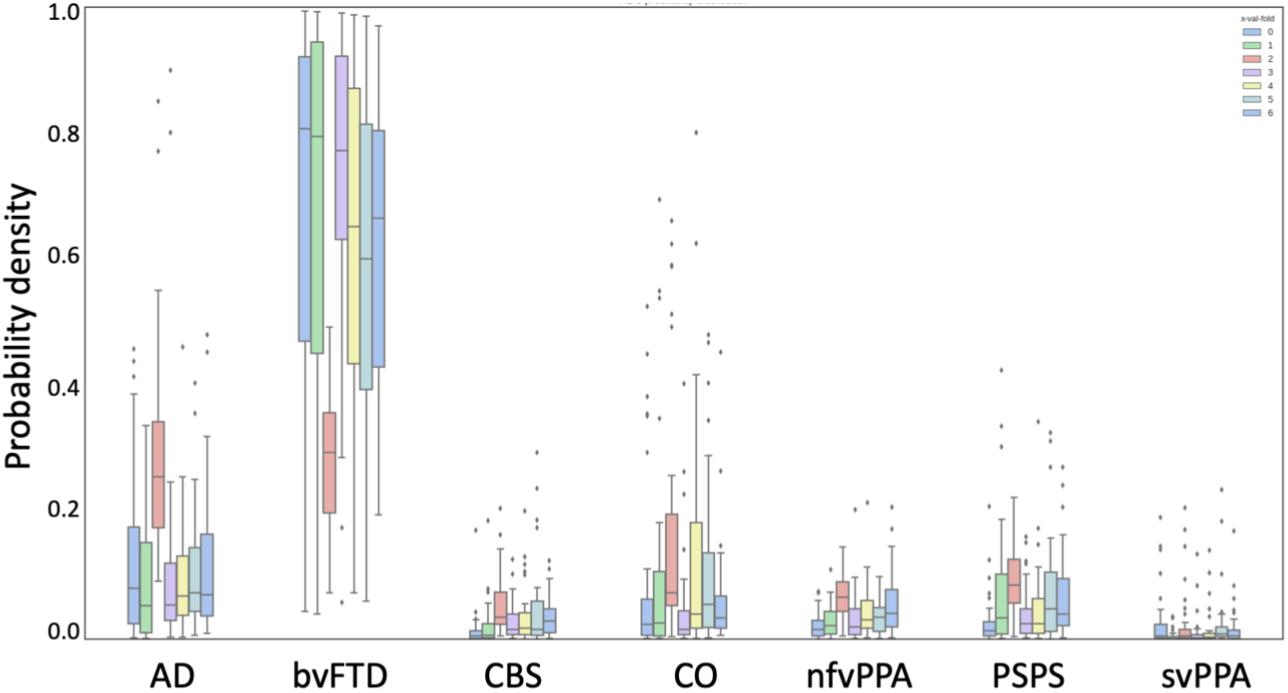
This is a bvFTD’s probability distribution using the model f2-20. The box plots represent the quartiles distributions for each outer-loop cross validation.

**Table 2:** Summary of the 10 best ranked multiclass models for all the dimension reduction (DR) models based on their true positive rate (TPR) and the *µ* index for the inner- and the outer-loop. The standard deviation (sd) is denoted between parentheses.

| Rank | Mask | DR | Inner-loop |  | <TPR> | p-value | Outer-loop |  |
| --- | --- | --- | --- | --- | --- | --- | --- | --- |
| | | | TPR(sd) | $\mu^*$ | | | TPR(sd) | $\mu^*$ |
| 1 | GM_WM | f2-20 | 0.76(0.23) | 7.20 | 0.73 | 0.71 | 0.71(0.25) | 7.20 |
| 2 | GM_WM | f4-0 | 0.80(0.21) | 5.07 | 0.73 | 0.29 | 0.65(0.27) | 5.07 |
| 3 | GM_WM | f8-0 | 0.79(0.23) | 6.30 | 0.73 | 0.42 | 0.66(0.27) | 6.30 |
| 4 | GM_MST | f2-0 | 0.78(0.24) | 6.59 | 0.72 | 0.43 | 0.66(0.28) | 6.59 |
| 5 | GM_MPST | Atlas-20 | 0.78(0.23) | 6.76 | 0.72 | 0.47 | 0.67(0.25) | 6.76 |
| 6 | GM_MST | Atlas-20 | 0.78(0.23) | 6.76 | 0.72 | 0.47 | 0.67(0.25) | 6.76 |
| 7 | GM_WM | Atlas-20 | 0.78(0.23) | 6.76 | 0.72 | 0.47 | 0.67(0.25) | 6.76 |
| 8 | GM_MPST | Atlas-15 | 0.77(0.22) | 6.76 | 0.72 | 0.52 | 0.68(0.26) | 6.76 |
| 9 | GM_MST | Atlas-15 | 0.77(0.22) | 6.76 | 0.72 | 0.52 | 0.68(0.26) | 6.76 |
| 10 | GM_WM | Atlas-15 | 0.77(0.22) | 6.76 | 0.72 | 0.52 | 0.68(0.26) | 6.76 |

\* $\times 10^{-2}$

**Table 3:** Pair-wise coupling, multiclass classification confusion matrix using the *f* 2 *−* 20 model in the GM WM mask. The results of the verification and validation set from the nested cross-validation using N subjects per fold.

|  | AD | bvFTD | CBS | CO | nfvPPA | PSP | svPPA | N |
| --- | --- | --- | --- | --- | --- | --- | --- | --- |
| Inner-loop |  |  |  |  |  |  |  |  |
| AD | 0.78(0.07) | 0.12(0.05) | 0.00(0.00) | 0.09(0.02) | 0.00(0.01) | 0.00(0.00) | 0.00(0.00) | 71 |
| bvFTD | 0.12(0.21) | 0.74(0.25) | 0.00(0.01) | 0.10(0.01) | 0.01(0.01) | 0.01(0.02) | 0.00(0.00) | 94 |
| CBS | 0.18(0.03) | 0.13(0.07) | 0.21(0.07) | 0.30(0.07) | 0.13(0.04) | 0.05(0.03) | 0.00(0.00) | 30 |
| CO | 0.02(0.02) | 0.02(0.01) | 0.00(0.00) | 0.95(0.02) | 0.00(0.00) | 0.00(0.00) | 0.00(0.00) | 124 |
| nfvPPA | 0.07(0.07) | 0.13(0.11) | 0.02(0.03) | 0.17(0.07) | 0.48(0.13) | 0.11(0.04) | 0.01(0.01) | 28 |
| PSP | 0.00(0.01) | 0.11(0.06) | 0.05(0.04) | 0.28(0.09) | 0.02(0.03) | 0.53(0.14) | 0.00(0.00) | 36 |
| svPPA | 0.06(0.05) | 0.13(0.11) | 0.00(0.00) | 0.00(0.00) | 0.02(0.02) | 0.00(0.00) | 0.79(0.10) | 26 |
| Outer-loop |  |  |  |  |  |  |  |  |
| AD | 0.70(0.05) | 0.13(0.10) | 0.01(0.03) | 0.14(0.08) | 0.00(0.00) | 0.01(0.03) | 0.00(0.00) | 14 |
| bvFTD | 0.10(0.13) | 0.67(0.20) | 0.00(0.00) | 0.15(0.05) | 0.06(0.10) | 0.02(0.03) | 0.00(0.00) | 18 |
| CBS | 0.17(0.14) | 0.13(0.15) | 0.23(0.24) | 0.33(0.19) | 0.10(0.15) | 0.03(0.07) | 0.00(0.00) | 5 |
| CO | 0.03(0.03) | 0.02(0.03) | 0.01(0.02) | 0.92(0.05) | 0.00(0.00) | 0.01(0.02) | 0.00(0.00) | 24 |
| nfvPPA | 0.10(0.10) | 0.23(0.18) | 0.00(0.00) | 0.30(0.10) | 0.27(0.15) | 0.10(0.10) | 0.00(0.00) | 5 |
| PSP | 0.00(0.00) | 0.19(0.16) | 0.02(0.05) | 0.26(0.10) | 0.02(0.05) | 0.50(0.11) | 0.00(0.00) | 7 |
| svPPA | 0.13(0.15) | 0.07(0.15) | 0.00(0.00) | 0.00(0.00) | 0.03(0.07) | 0.00(0.00) | 0.77(0.23) | 5 |

**Table 4:**
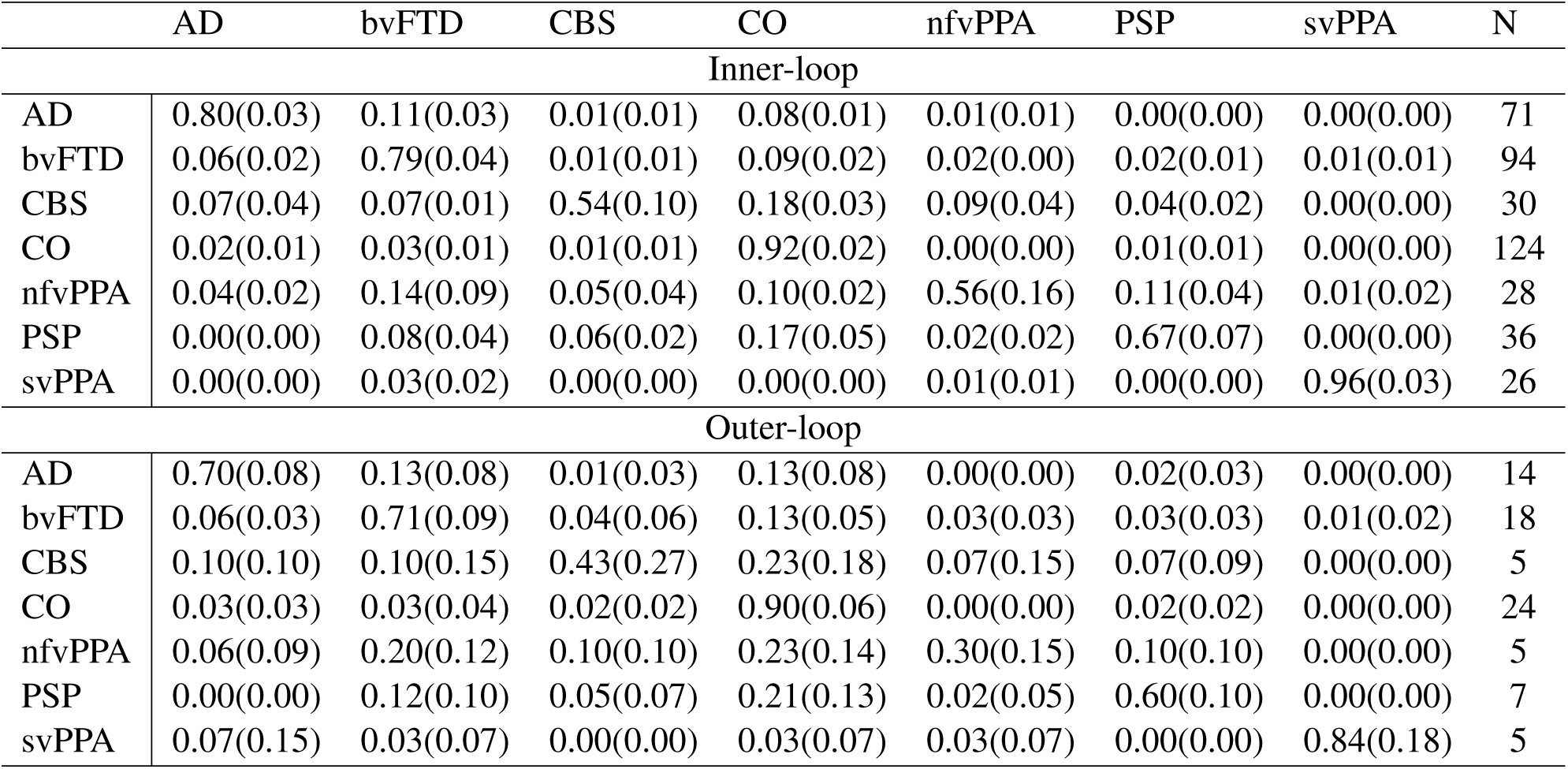
Confusion matrices comparing the efficiency of probability interpretation schemes. Pair-wise coupling, multiclass classification confusion matrix using the *f* 2 *−* 20 model in the GM WM mask and corrected using the logistic regression. The results of the verification and validation set from the nested cross-validation using N subjects per fold.

### W-maps, clinical, and genetic results explain misclassifications

Figure 3 shows the value of the lowest mean W-score of the brain for each case in the entire cohort. The majority of misclassifications of disease groups as CO occurred in cases where the maximum atrophy was within the w-score range of maximal atrophy in CO. From the 84 cases clinically diagnosed with AD, 11 cases (13%) were misclassified as CO. The majority of these misclassified AD cases (9 cases -- 81%) were early-age-of-onset AD (EOAD) patients. This suggests that the larger brain volume in EOAD patients leads to misclassifications and that larger cohorts of EOAD are needed to optimize algorithm performance in EOAD. EOAD cases have not been included in previous machine learning classification algorithms. From the 108 cases clinically diagnosed with bvFTD, 14 cases (14%) were misclassified as CO. Half the misclassified bvFTD cases were genetic: *C9orf72* mutation in 6 cases (43%) and one MAPT mutation (7%)). Two patients had FTD phenocopy,[38] one had argylophilic grain disease pathology and three had no volume loss on the MRI and met only possible bvFTD criteria [62]. From the 30 cases clinically diagnosed with CBS, 7 cases (23%) were misclassified as CO and all of these cases had minimal volume loss in the lateral parietal region (their region of largest volume loss). From the 30 cases clinically diagnosed with nfvPPA, 7 cases (23%) were misclassified as CO. All cases had minimal volume loss in the left perisylvian region. From the 42 cases clinically diagnosed with PSPS, 10 cases (23%) were misclassified as CO. Six of the misclassified PSPS cases showed midbrain volume loss and no gross cortical atrophy and the remaining 4 cases showed minimal gross atrophy with no gross midbrain atrophy. From the 30 cases clinically diagnosed with svPPA, 1 case (3%) was misclassified as CO despite having gross left anterior temporal volume loss. From the 144 cases clinically diagnosed as CO, 15 cases (10%) were misclassified as having a neurodegenerative disease (4 AD, 5 bvFTD, 3 CBS, and 3 PSPS). The majority of these cases had minimal bilateral lateral parietal volume loss. Five clinically normal CO had a positive genetic mutation and were all classified as CO (three were *C9orf72* carriers and 2 were PRGN carriers). None of the misclassified CBS, nfvPPA or PSPS cases had genetic mutations.

**Figure 2:**
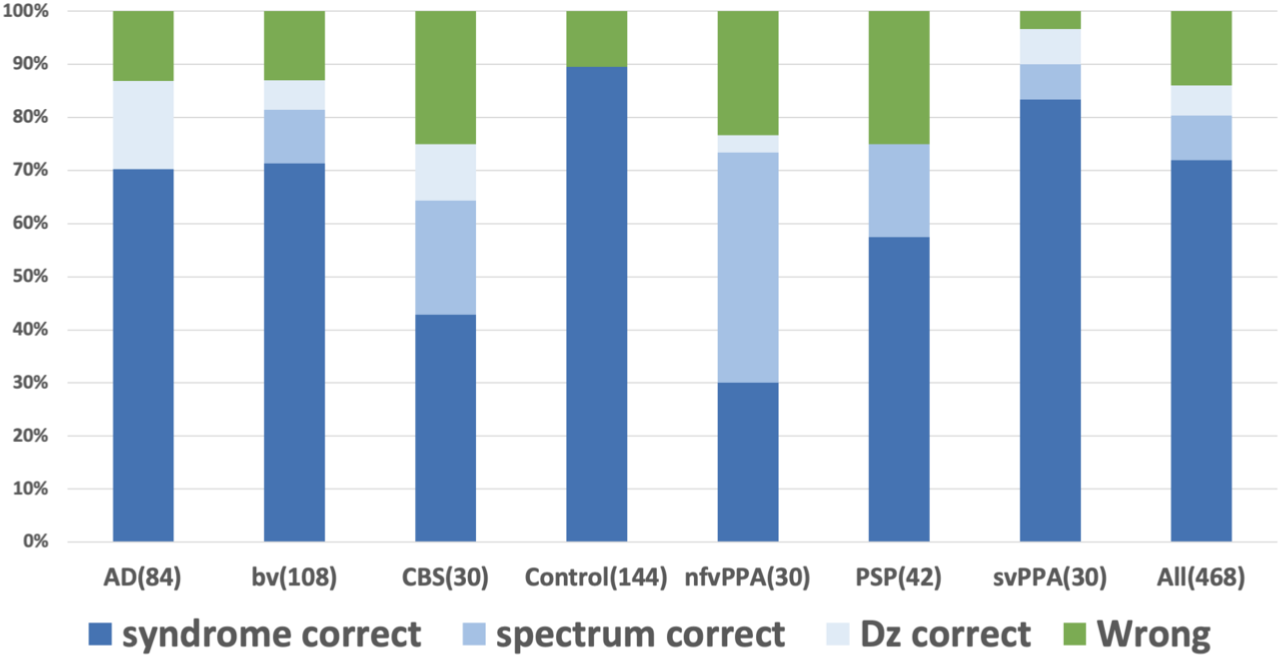
RVM performance against clinical diagnosis.

**Figure 3:**
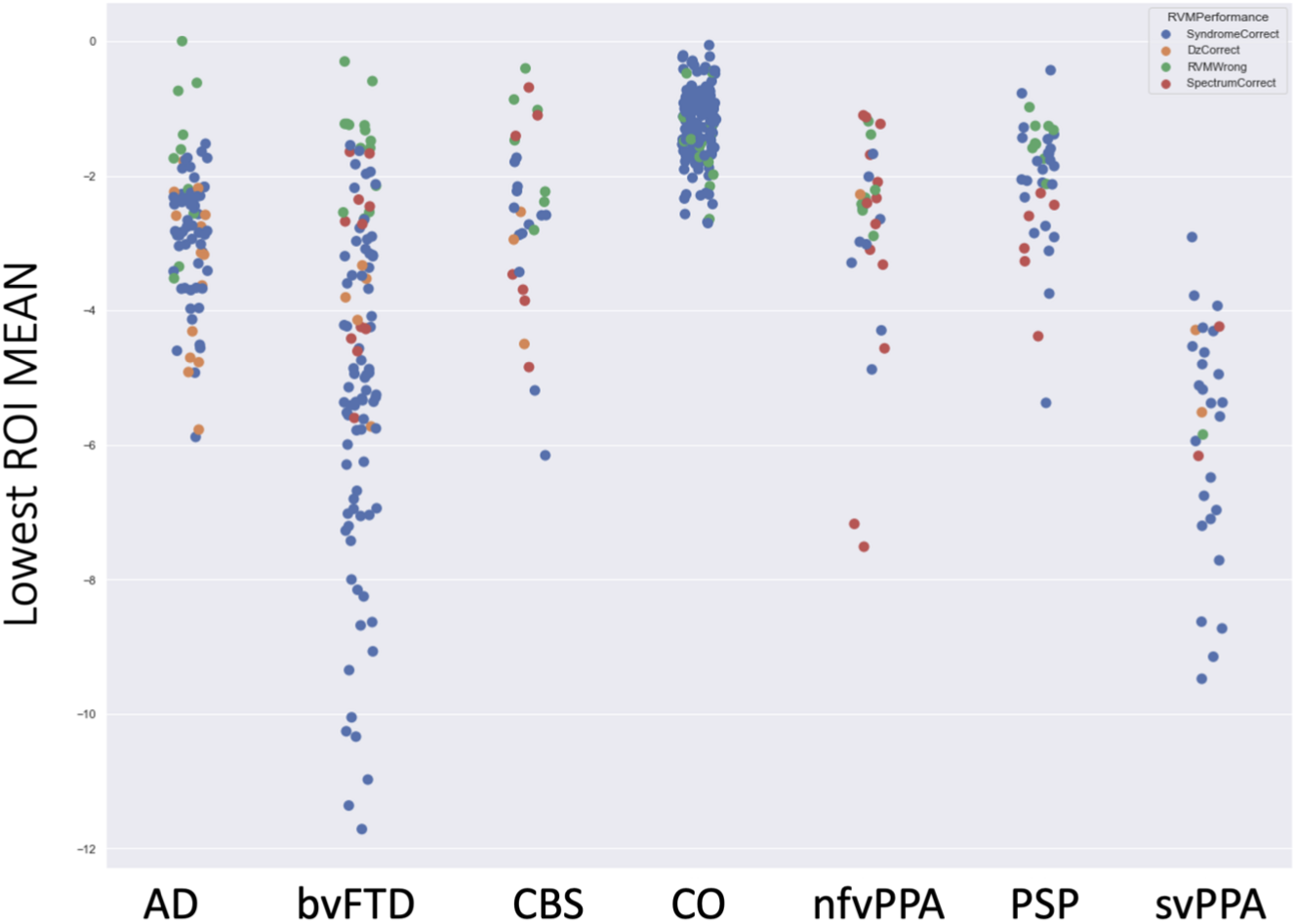
RVM performance against W-score maps.

### Pathological results explain misclassifications

Figure 4 and Table 5 show RVM performance in the 72 cases with pathological diagnosis. Notably, when misclassifications happened, RVM identified a neurodegenerative process in 89% of the cases. Of 21 cases misclassified, 29% had atypical pathology for that syndrome, 33% were PSP, 24% were taupathies other than PSP (CBD and Pick’s), and 14% were TDP pathology (1 FTLD-TDP-A, 1 FTLD-TDP-B, 1 FTLD-TDP-C). Eight out of 72 (11%) of the cases with neurodegenerative process were misclassified as CO and five of these cases were PSPS.

**Figure 4:**
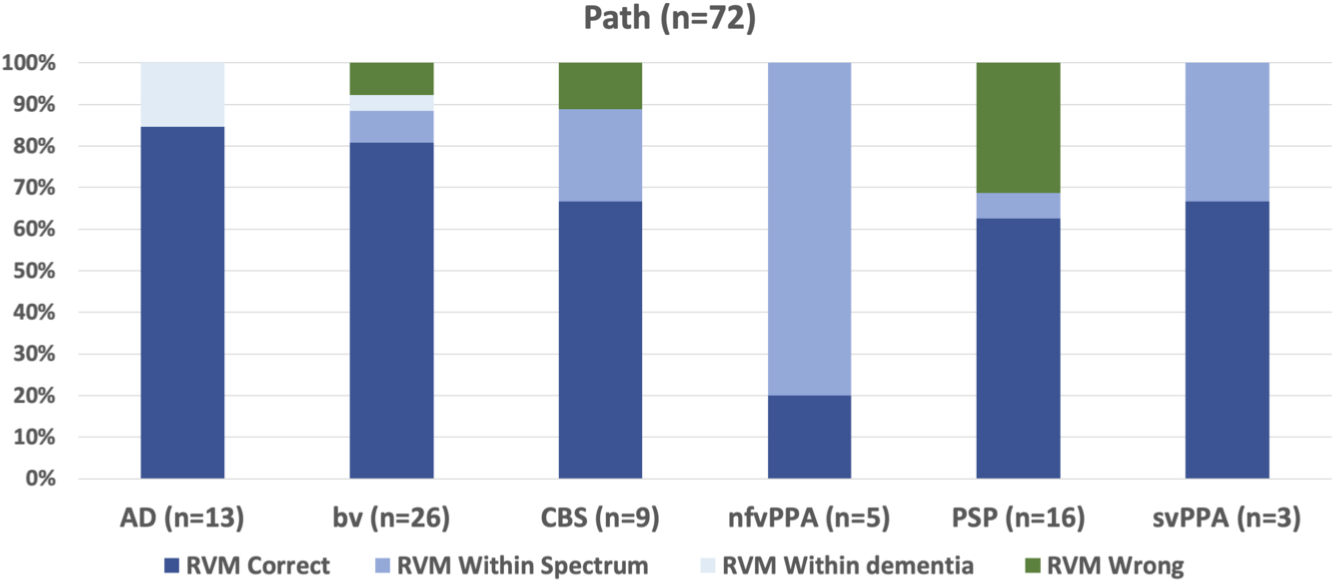
RVM performance against pathological diagnosis.

**Table 5:**
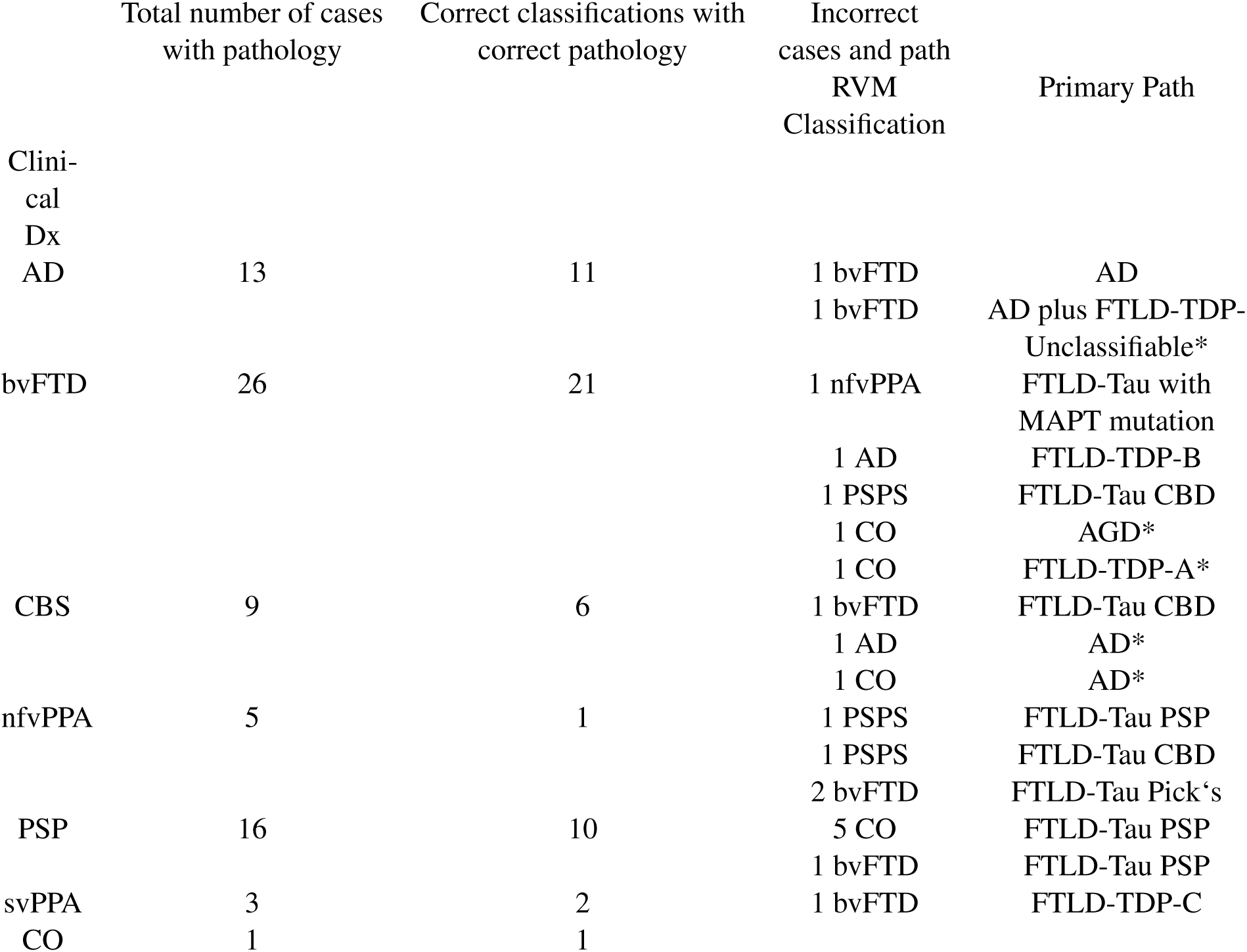
RVM performance in in comparison to pathological diagnosis.

Thirteen cases were diagnosed clinically as AD and had pathological data, all had Alzheimer’s disease as the primary pathology, two were misclassified by RVM as bvFTD and the remaining 11 were correctly classified as AD. One of the misclassified cases had FTLD-TDP Unclassifiable as a primary secondary pathology.

Twenty-six cases were diagnosed clinically as bvFTD and had pathological data, 26 had FTLD pathology as the primary pathology. Two cases were misclassified by RVM as CO, one as AD, one as nfvPPA, one as PSPS, and the remaining 21 were correctly classified as bvFTD. The cases misclassfied as CO were Argylophilic grain disease and FTLD-TDP type A. The details of the remaining diagnostic categories are shown in TableXX.

## Discussion

### Summary of the main findings

In this study, we implemented RVM and logistic classification on whole-brain T1-weighted MRI tissue segmentation data in a multi-class classification approach aiming to distinguish AD, CO, and the five canonical FTD-spectrum syndromes: bvFTD, CBS, nfvPPA, PSPS and svPPA. The algorithm, in the context of multi-class classification, showed high performance for classifying CO, moderate to high performance diagnosing AD, bvFTD, and svPPA, and marginal (i.e. a little better than chance, 14%) to modest performance diagnosing nfvPPA, PSPS, and CBS. Among the misclassified cases, there were higher percentages of EOAD in AD and genetic bvFTD. Even in cases where the algorithm did not identify the precise syndrome, it was usually able to differentiate AD from non-AD pathology, and cases from controls. Examining MRI W-scores revealed that the cases that were misclassified as controls tended to have minimal atrophy. While these findings reinforce the challenges associated with MRI-based classification of these heterogenous neurodegenerative disorders, they also highlight potential uses of even imperfect classification systems and point toward opportunities to improve imaging-based diagnosis over time.

### The advantages of our study

Strengths of our study included the implementation of RVM with logistic classification where knowledge from the inner-loop was used to correct the diagnosis probability distribution, the incorporation of all clinical syndromes in a large neuroimaging cohort, the use of a single, whole brain MRI alone as input, and the comparison of the probability outcome against objective disease markers including genetic mutation status, pathological data and quantitative MRI measures. This process allowed us to explore and understand the potential reasons for misclassifications occurring with our method. In our study, RVM and logistic classification produced higher accuracy than reported in the literature using SVM [24, 25, 22]. We used a whole brain tissue segmentation approach which is data-driven and more sensitive to the considerable spatial variability of brain changes in neuropsychiatric diseases [9, 63]. In agreement with the literature, we found a higher accuracy by incorporating white matter tissue measures. However in our study both gray and white matter masks were extracted from one MRI sequence, T1-weighted, without incorporating other modalities like diffusion tensor imaging (DTI) or FLAIR sequences [24, 18]. Clinically, the probability of a diagnosis is specific within each differential diagnosis. For instance, when both AD and svPPA features are considered, the mere presence of svPPA probability above random choice might strongly sway a physician to consider it as the primary diagnosis - given the high specificity of its neuroimaging and clinical characteristics - even though AD is naturally more prevalent [64, 4]. Taking into account that certain specific features carry higher diagnostic weight than others, we attempted to emulate this phenomena by implementing logistic classification on the entire probability distribution provided by RVM. In this mode, each case simultaneously has a probability estimate of each of the possible diagnostic classes. For instance, if the continuous probability distribution for a case is (AD: 26%, bvFTD: 18%, CBS: 2%, CO: 23%, nfvPPA: 4%, PSPS: 1% and svPPA: 24%), the RVM winner is AD because it has the highest probability. However, this approach ignores the rest of the information from the probability distribution. Given the unique specificity for some imaging features and the unequal proportion of disease prevalence, a 24% probability for svPPA carries a higher degree of confidence than 26% probability for AD. There, we alleviated these types of errors by implementing the posterior logistic classification approach that takes into account the whole probability distribution rather than the highest probability alone.

### Why its difficult to classify neurodegenerative illnesses

Neurodegenerative diseases target vulnerable brain networks. Brain atrophy starts focally and spreads over the course of years to involve highly connected yet distant brain regions [65, 66, 67]. Consequently, the clinical syndrome is dictated by the neuroanatomical networks involved [68, 69, 70]. MRI can aid the diagnosis of neurodegenerative diseases and to some extent help predict the underlying neuropathology [68, 71]. Although bvFTD is associated with prefrontal and temporal atrophy with the anterior insula, cingulate, orbitofrontal, dorsolateral prefrontal and anterior temporal regions being more commonly implicated [72], a specific bvFTD molecular pathology, TDP-43 type B, -both in sporadic and *C9orf72* repeat expansion cases -, is known to not cause overt MRI brain atrophy [72]. Further, FTD phenocopy and patients meeting only possible bvFTD criteria show no overt cortical atrophy [35]. Finally, rare pathologies such as argyrophilic grain disease and globular glial tauopathies can be included in FTD cohorts. Finally, rare pathologies such as argyrophilic grain disease and globular glial tauopathies can be included in FTD cohorts.

Lacking large sample sizes and well-defined imaging characteristics contribute to worsen specificity. In our study, bvFTD misclassifications happened when there was no significant cortical atrophy. Half the misclassified bvFTD cases were genetic: *C9orf72* mutation in six cases representing 43% of the bvFTD cases and one MAPT mutation representing 7%. Two patients had FTD phenocopy, one had argylophilic grain disease pathology and three met only possible bvFTD criteria. All the misclassified AD cases had minimal amount of volume loss and the majority of these were EOAD cases. EOAD cases have not been included in previous automated classification studies. Our findings suggest that the inclusion of larger sample sizes of atypical and early forms of AD is needed in order to achieve generalizable machine learning algorithm results.

Our algorithm exhibited marginal to modest performances with PSPS, CBS, and nfvPPA. These low accuracies reflect the complexity and difficulty to diagnose those illnesses potentially given the relatively small sample size for these more rare diagnoses, in contrast to bvFTD and PPA where neuroimaging increases diagnostic specificity [7, 73]. Although predominant midbrain atrophy on MRI or hypometabolism on positron emission tomography (PET) in PSPS [74] and discernible frontal atrophy on brain MRI have been reported in 60% of PSP-Richerdson syndrome patients [75, 76, 77], they are considered only supportive features in the current diagnostic criteria [74]. Similarly, a few studies suggest that CBS patients have volumetric changes in premotor cortices, supplemental motor area, and insula [78, 79]. These findings are inconclusive and as a result are not considered in the current CBS diagnostic criteria [80]. It is also important to note that MRI was found to exhibit low sensitivity in nfvPPA diagnosis [81].

Interestingly, the healthy control group contained a few patients who turned out to be FTD genetic mutations carriers. These cases were classified as CO by RVM, and this finding points to the importance of genetic data when choosing a control cohort. Given the known MRI changes in presymptomatic genetic carriers, [82] it is unclear how the inclusion of such cases in the control group might influence algorithm classification accuracy. Further, the seven healthy controls misclassified as AD or CBS showed volume loss in the bilateral lateral parietal regions which are commonly targeted in AD or CBS. These findings highlight the variability within the control sample and the role of cognitive resilience in patients who are asymptomatic and perform well on cognitive testing despite having brain volume changes [83]. We can not clearly explain the reason for the misclassification of eight healthy controls as bvFTD or PSPS as these cases did not show clear atrophy. We speculate that this might be because of the clinical PSPS and bvFTD cases that did not exhibit cortical atrophy and were included in the training set of the respective disease cohort. Brain MRIs, for cases like these, represent a challenge for classification algorithms. Previous studies incorporated a small to modest cohort of controls [24, 25, 18, 22, 23], considered subjective cognitive impairment patients as controls [19, 20], or did not include a control group [21]. Our study emphasizes the importance of including a healthy control cohort when developing a differential diagnostics tool. We show a high accuracy (86%) distinguishing healthy subjects from patients with any neurodegenerative disease using structural MRI.

### Comparison to other machine learning classification studies

In the machine learning classification studies based on the most widely available and the least technically demanding MRI imaging approach, *e.g.* whole brain T1-weighted, binary classification algorithms have demonstrated accuracies that ranged: between 76% and 95% for differentiating AD from CO [10, öppel2008**?**, 84, 85, 86], between 77% and 85% in differentiating FTD from CO [87, 88] and between 70% and 85% in differentiating AD from FTD [9, 87, öppel2015**?**]. Several studies attempted a multi-class classification on various cohort with fluctuating accuracies. Multi-class classification studies showed accuracies ranging between 50% and 76% in different cohorts including AD, FTD, LBD, and vascular dementia [öppel2015**?**, 19, 89, 90, 18, öller2016**?**, 9]. Although, the outcome measured and the type of neurodegenerative diseases varied across studies, our study showed either higher diagnostic accuracy or a more inclusive dataset compared to other studies. Differentiating FTD, from AD and CO, Bron et al. reported 0.85 area under the curve using VBM-GM whole brain analysis [24]. Dukart et al. reported 73% accuracy using GM whole brain analysis [25]. Kuceyeski et al. reported 84% accuracy using a combination of GM and DTI [18]. None of these studies distinguished within the FTD spectrum. Koikkalainen et al. and Tong et al. implemented a multi-class State Index Classifier to study the same sample of AD, vascular dementia, FTD, LBD and subjective cognitive impairment patients. They reported up to 59% accuracy. Although, as pointed by the authors, the LBD cases might be lowering the accuracy, these studies did not distinguish among the FTD variants and did not include a cohort of healthy controls [19, 20]. Kloppel et al. prospectively studied AD, FTD, LBD, and CO and reported area under the curve of 0.76 for AD and 0.78 for FTD. However, small numbers of FTD (18 subjects) and LBD (4 subjects) were included, and FTD was not separated into its variants [22]. Vemuri et al. in the only multi-class study that includes pathological data reported sensitivities about 91%, 79%, and 84% for AD, LBD, and FTLD-TDP respectively. However, the study included only cases with pure pathology and excluded FTLD cases with pathologies other than FTLD-TDP [23], including FTLD-tau could have potentially reduced these accuracies given the added heterogeniety.

Our study is consistent with the literature and it strives to explain the reasons for misclassification relying upon clinical, genetic, and pathological evidences. We found that patients with minimal atrophy, genetic mutations, FTD phenocopy, and EOAD are less likely to be classified correctly. The most common misclassification is the minimal brain volume loss. (Joie 2012, Ossenkoppele 2015) which in specific disease etiologies is known as they present without causing significant MRI changes. [72, 35] Those findings suggest, based on the analysis of the pathological and genetic data of this cohort, that our algorithm converges to similar difficulties faced by dementia specialists.

This study does not include LBD. LBD tends to have a lower diagnostic accuracy even with implementing the clinical criteria [91]. The sparsity of LBD in our available bank of diseases did not allow us to include this dementia in our cohorts, and we acknowledge the limitation in our findings caused by this missing important category. Inherently to the design of our study, we included patients younger than 70 years of age because the diagnostic challenge is more relevant in relatively younger patients when the prevalence of AD and FTD is comparable [37]. However, including all ages in algorithm training might leads to inaccuracies as AD is more predominant in older patients. Lastly, MRI images included in this study requires processing which is a limitation to many current machine learning techniques.

## Conclusion

Our study provides further evidence that incorporating automatic approaches into clinical practice can be an efficient and economical way to provide services to the increasing number of patients with cognitive symptoms worldwide. Machine learning algorithms are capable of providing valuable help that is close to a dementia specialist opinion, and can lead to rapid and early triage and consequently appropriate therapeutic care and interventions, if available, to the increasing numbers of patients with neurodegenerative disease. The goal of such algorithms is likely to be more relevant outside dementia centers where physicians and radiologists typically have less experience with dementia diagnostics. Our findings indicate that misclassifications are secondary to limitations related to the complexity of neurodegenerative illnesses and argues that incorporating larger datasets will improve the accuracy statistics. This also suggests that misclassifications are an integral part of attempting to diagnose neurodegenerative diseases and it can be argued that a margin of error is acceptable as it highlights the risk group which can benefit from a tertiary care evaluation and potentially further testing. Finally, this study suggests that incorporating detailed phenotyping, pathological, and genetic data into machine learning studies can provide clues for the interpretation of machine learning outputs which in turn can lead to better algorithm inferential reproducibility.

## Data Availability

All data produced in the present study are available upon reasonable request to the authors

## Acknowlegment

Quest Diagnostics.

Brainsight.

## Abbreviations

AD: Alzheimer’s disease
bvFTD: behavioral variant frontotemporal dementia
CBS: cortibobasal syndrome
CBD: corticobasal degeneration
CO: Controls
FTLD: frontotemporal lobar degeneration
nfvPPA: non-fluent variant primary progressive aphasia
PSPS: progressive supranuclear palsy syndrome
RVM: relevance vetcor machine
svPPA: semantic variant primary progressive aphasia.

## Footnotes

1 https://www.siemens-healthineers.com/

2 http://www.fil.ion.ucl.ac.uk/spm

3 http://www.neuromorphometrics.com/

